# Spatial transcriptomics reveals layered immune-metabolic architecture of giant cell myocarditis

**DOI:** 10.64898/2025.12.15.25342188

**Authors:** Nicolas Musigk, Alexander Sudy, Phillip Suwalski, Katharina Jechow, Johannes Liebig, Philip Bischoff, Isabell Anna Just, Volkmar Falk, Felix Schoenrath, Roland Eils, Ulf Landmesser, Christoph Knosalla, Christian Conrad, Bettina Heidecker

**Author notes:** Corresponding Author: Prof. Dr. Bettina Heidecker, MD, FESC, FACC, Deutsches Herzzentrum der Charité, Campus Benjamin Franklin, Department of Cardiology, Angiology and Intensive Care Medicine. Shared First-Authorship (Nicolas Musigk, Alexander Sudy, Phillip Suwalski). Shared Last-Authorship (Christian Conrad, Bettina Heidecker).

## Abstract

Giant cell myocarditis (GCM) represents the most fulminant form of inflammatory cardiomyopathy, yet its spatial pathophysiology remains poorly defined. We applied full-transcriptome spatial capture in-situ RNA sequencing to a human GCM explant, generating a comprehensive high-resolution molecular atlas across twelve cardiac localizations. Spatial clustering and cell-type deconvolution delineated a complex cellular landscape dominated by cardiomyocytes, fibroblasts, and immune cells, with pronounced transcriptional heterogeneity along the epicardial-endocardial axis. Myeloid cells localized to inflammatory hotspots and exhibited *SPP1*- and *IL1B*-driven activation, whereas lymphoid cells displayed a continuum from IgM- to IgG4-secreting plasma-cell differentiation. Layer-resolved pathway analysis revealed epicardial enrichment of IL-6/JAK/STAT3 and TNF-α/NF-κB signaling, myocardial upregulation of oxidative phosphorylation and myogenesis, and endocardial activation of stress and apoptosis programs. These data uncover a layered immune-metabolic architecture linking epicardial inflammation to myocardial remodeling and endocardial stress, providing a spatial framework for understanding immune-mediated myocardial injury in GCM.

## Introduction

As of today, giant cell myocarditis (GCM) exhibits the most severe and clinically devastating trajectory within the spectrum of inflammatory myopericardial syndromes (IMPS). As the most fulminant inflammatory heart disease, GCM frequently progresses to advanced heart failure within a few months of diagnosis^1^. Many patients ultimately require mechanical circulatory support or heart transplantation (HTx), despite receiving aggressive multi-agent immunosuppressive therapy^1–3^.

Notwithstanding the urgent need for better therapies, the pathophysiology of GCM and its spatial dynamics of inflammation remain poorly understood. Current paradigms of myocarditis pathogenesis, as highlighted in the 2025 European Society of Cardiology Guidelines on IMPS, suggest that inflammation frequently involves the pericardium and myocardium simultaneously^3^. The role of serous layers in initiating inflammation is established in other systems such as the pleura or the meningeal membranes, underscoring the understanding that inflammation is not strictly compartmentalized but can propagate across contiguous tissue planes^4,5^. By analogy, this raises the conceptual possibility of a similar “outside-in” trajectory in IMPS, in which inflammation could extend inward from the pericardial interface toward the myocardium. At present, this notion remains entirely speculative, owing to the rarity of tissue samples and the limited spatial resolution of bulk transcriptomic and standard histologic approaches.

However, GCM may offer a unique window to explore this newly hypothesized “outside-in” concept, as its pronounced and destructive inflammatory response may amplify spatial patterns that are otherwise difficult to resolve in milder myocarditis forms. By applying spatial transcriptomics to an explanted human GCM heart, we aimed to map the distribution of immune and stromal cells across cardiac tissue layers and to characterize transcriptional programs along the epicardial-endocardial axis. Here, we demonstrate that spatially resolved gene expression profiling of GCM captures the profound immune activity characteristic of this disease and reveals spatial patterns compatible with, and suggestive of, a potential pericardium-to-myocardium progression of inflammation in myocarditis.

## Results

### Cellular landscape of the GCM-affected heart

We performed full transcriptome, spatially resolved RNA sequencing using Stereo-seq (STOmics) on 20 samples covering 12 distinct cardiac localizations (Fig. 1). After binning 100 × 100 spots (corresponding to 50 × 50 µm areas) and applying spatially aware unsupervised clustering, we generated high-resolution spatial gene-expression maps of the tissue sections (Fig. 2a). This approach captured all major cardiac cell types including cardiomyocytes, fibroblasts, and adipocytes and revealed their characteristic spatial organization within the heart wall, with quantification across samples demonstrating consistent representation of these populations (Fig. 2b-c). Hierarchical clustering of the top differentially expressed genes per cluster confirmed the transcriptional identity of the major cell types (Fig. 2d). Spatially resolved clustering uncovered a complex cellular landscape dominated by cardiomyocytes, adipocytes, and stromal cells, predominantly fibroblasts, together with smaller endothelial, myeloid and lymphoid cell fractions.

**Figure 1:**
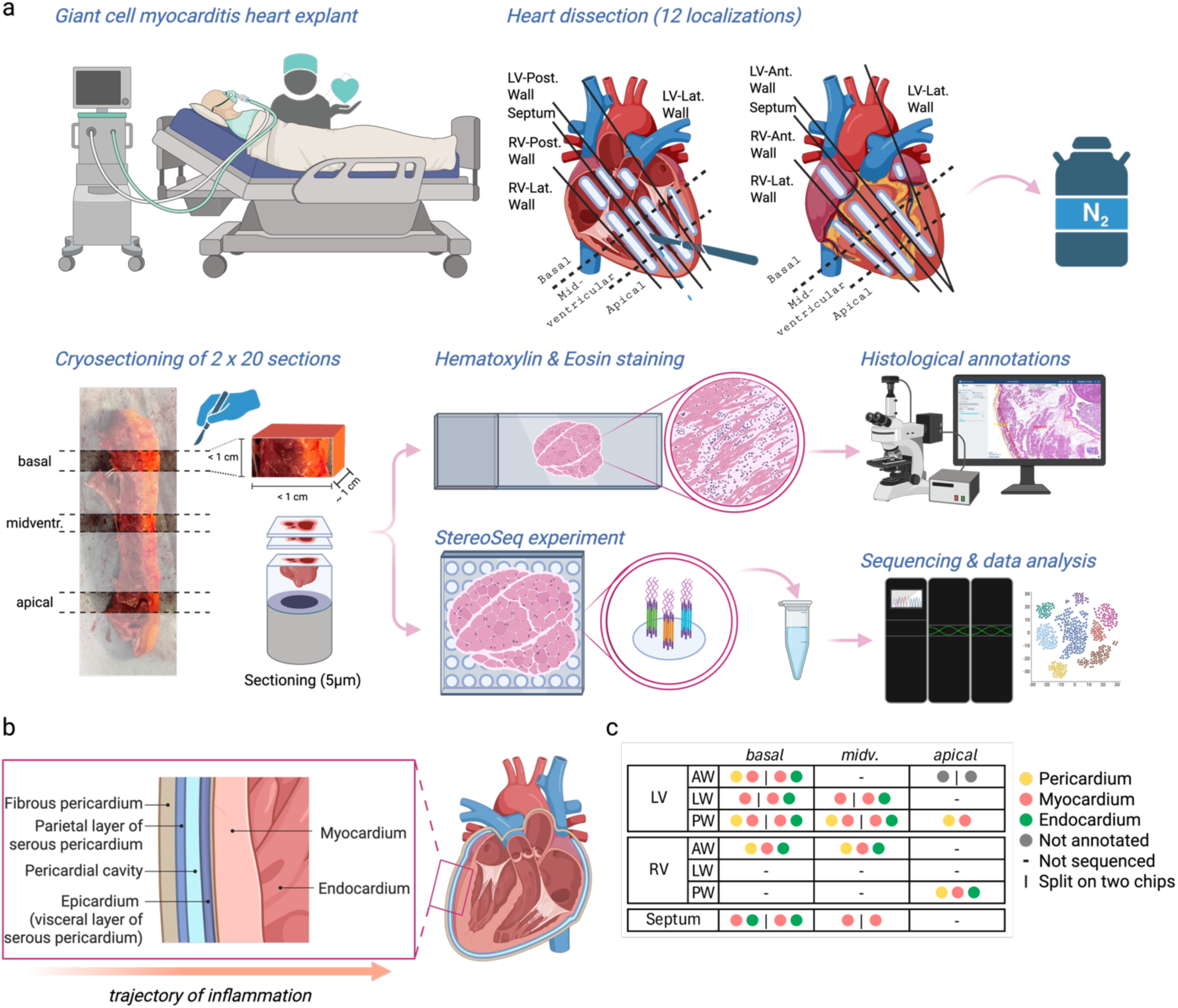
Overview of the experimental setup and sample processing. (a) Workflow overview - from bedside to bench. Following heart explantation, the heart was dissected into strip-like pieces and flash-frozen in liquid-nitrogen-cooled isopentane. From these, 1 × 1 × 1 cm blocks were extracted from basal, midventricular, and apical regions and embedded in optimal cutting temperature compound for cryo-sectioning. The resulting sections were subjected to Stereo-seq analysis. (b) Schematic representation of the anatomical structure of the ventricular wall. (c) Overview of the tissue localizations included in the Stereo-seq experiments.

**Figure 2:**
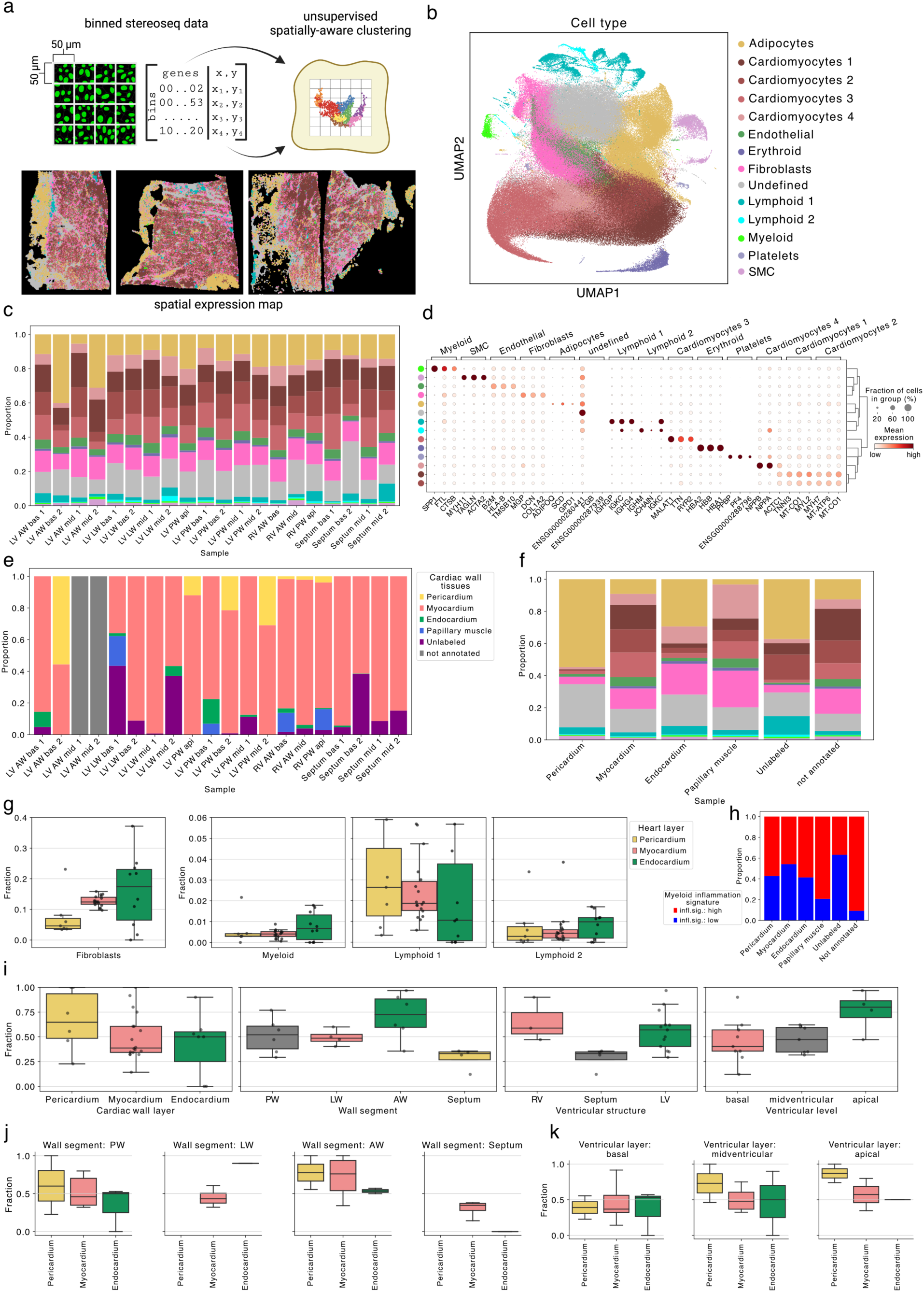
Unsupervised spatially aware clustering identifies major cardiac cell types and maps spatial heterogeneity in cell-type abundance and myeloid activation across 12 cardiac regions. (a) Overview of processing steps. SpatialLeiden clustering was performed on binned Stereo-seq data (100 × 100 spots = 50 × 50 µm) to generate spatial gene-expression maps, three of which are shown as examples. (b) Uniform Manifold Approximation and Projection (UMAP) of all 50 × 50 µm Stereo-seq bins (n = 310,150). (c) Relative abundances of identified cell types across all samples. Colors correspond to clusters in (b). (d) Hierarchical clustering of the top three differentially expressed genes per cell type (Wilcoxon rank-sum test). Colors correspond to clusters in (b). (e) Relative proportions of cardiac wall layers across all Stereo-seq experiments. (f) Cell-type composition of cardiac wall layers and papillary muscles. (g) Relative abundances of immune-related cell types within cardiac wall layers and papillary muscles. (h) Comparison of highly versus lowly activated myeloid cells. (i) Spatial distribution of myeloid activation across all sampled cardiac localizations. (j,k) Myeloid activation across wall segments (j) and ventricular levels (k) grouped by cardiac wall layer.

All cardiomyocyte populations expressed canonical contractile genes (*TNNT2, MYH6, ACTN2, TTN, RYR2*), confirming their identity. However, spatially resolved clustering revealed four transcriptionally distinct cardiomyocyte states, reflecting metabolic and structural heterogeneity within the myocardium. These states were classified as *Cardiomyocyte 1-4*. *Cardiomyocyte 1* exhibited increased expression of *TNNI3, MT-CO1*, and *MYL2*, whereas *Cardiomyocyte 2* showed elevated levels of *MYH7, MT-ATP6,* and *MT-CO1*. Both clusters thus displayed enhanced sarcomeric and mitochondrial gene programs consistent with metabolic maturation, increased contractile capacity, and remodeling as seen in both adaptive and pathological hypertrophy^6–12^. *Cardiomyocyte 3* exhibited a metabolic and structural gene program characterized by elevated expression of *MALAT1, TTN,* and *RYR2*, indicative of pathological remodeling, altered calcium handling, and increased apoptotic susceptibility^13–15^. *Cardiomyocyte 4* upregulated *NPPA, NPPB,* and *ACTC1*, defining a molecular signature consistent with cardiac stress responses^16–19^. These transcriptional gradients suggest that cardiomyocytes in GCM could undergo distinct metabolic and stress-response adaptations within the inflamed myocardium.

The organization of lymphoid cells into two distinct clusters may reflect the presence of functionally and developmentally distinct B cell populations within the inflamed cardiac wall in GCM. *Lymphoid 1* expressed immunoglobulin-related genes, including *IGHG4*, *IGKC*, and *IGHGP,* consistent with mature B cells and plasma cells that have undergone class-switch recombination and are actively producing IgG4 antibodies. This pattern reflects advanced B cell differentiation and isotype switching driven by chronic antigen stimulation and T follicular helper cell interactions, as described in IgG4-related disease and other chronic inflammatory states^20,21^. The predominance of IgG4-secreting cells within tissue is a recognized feature of chronic immune activation and may contribute to local immune regulation or pathology^21,22^. *Lymphoid 2* was characterized by expression of *IGHM, JCHAIN,* and *IGKC,* representing an earlier stage of B cell activation with IgM secretion and joining chain expression required for multimeric IgM and IgA formation. This population corresponds to naïve or early memory B cells responding to acute antigenic stimulation prior to class-switch recombination and full plasma-cell differentiation^23–25^. Together, these two populations may delineate a continuum of B cell maturation-from early IgM-secreting cells to class-switched IgG4-secreting plasma cells-mirroring germinal-center-like activity and adaptive humoral responses within the inflamed cardiac wall. These transcriptional gradients suggest that B cells in GCM could undergo metabolic and stress-response adaptations, consistent with active antibody production and a dynamic, heterogeneous humoral immune milieu.

Together, these data establish a comprehensive reference map of the cellular composition, transcriptional diversity, and spatial organization of GCM.

### Spatial heterogeneity of gene expression and immune cell activity in GCM

To investigate spatial heterogeneity of gene expression and immune cell activity in GCM, we analyzed cell-type composition and myeloid activation across 12 cardiac localizations. By integrating Stereo-seq–derived transcriptomic profiles with anatomical annotation of cardiac wall layers, we identified marked differences in cellular abundance and activation states along the epicardial–endocardial axis (Fig. 2e–k). Pericardial sections were enriched in immune and stromal cell populations, including fibroblasts, lymphoid, and myeloid cells, whereas cardiomyocytes predominated within the myocardium and endocardium (Fig. 2e-g). Quantitative assessment of myeloid activation suggests a spatial gradient, with the highest fraction of highly activated myeloid cells toward the pericardial side, gradually decreasing across the myocardial wall (Fig. 2h–k). This pattern was consistent across ventricular levels and wall segments, supporting the potential presence of a compartmentalized inflammatory architecture with epicardial immune activation and progressive attenuation toward the endocardium.

### Spatially resolved pathway enrichment across cardiac layers

To delineate molecular differences across cardiac wall layers, we performed gene set enrichment analysis (GSEA) using the MSigDB hallmark gene sets^26^. Distinct enrichment and depletion patterns were observed across the pericardial, myocardial, and endocardial layers, spanning inflammatory, signaling, metabolic, and stress-response pathways (Fig. 3). The pericardium displayed the strongest enrichment of immune and inflammatory pathways, such as IL-6/JAK/STAT3 and TNF-α/NF-κB signaling, suggesting localized cytokine activation at the epicardial interface. In contrast, myocardial regions showed relative depletion of inflammatory programs but enrichment of oxidative phosphorylation (OXPHOS), MYC targets V1, and myogenesis pathways, consistent with metabolic adaptation and structural remodeling in response to chronic inflammation. Endocardial sections displayed increased expression of apoptosis, hypoxia, and unfolded protein response pathways, indicative of enhanced cellular stress and damage signaling in deeper myocardial layers.

**Figure 3:**
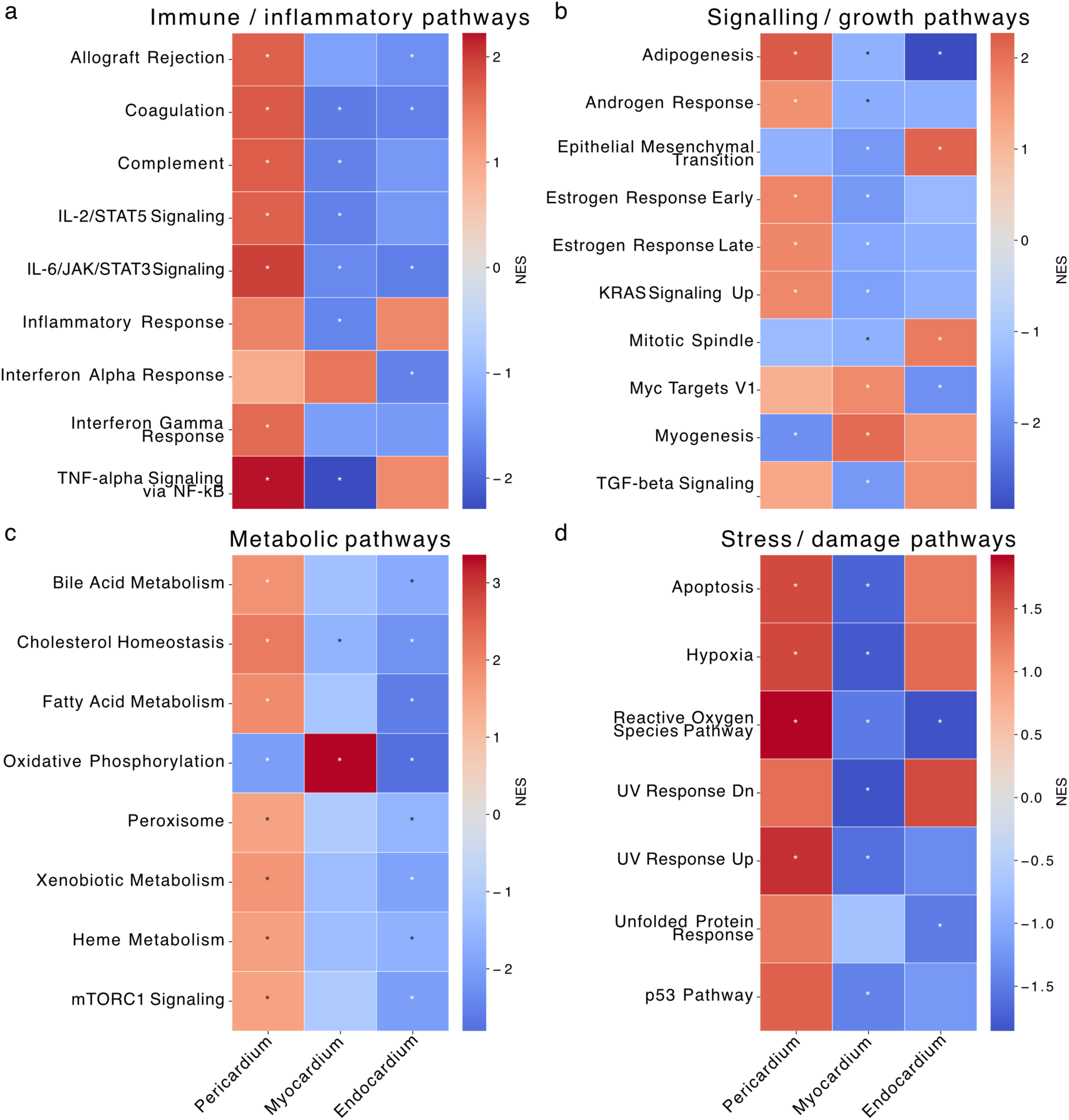
Gene set enrichment analysis of MSigDB hallmark pathways across cardiac wall layers. Gene set enrichment analysis (GSEA) was performed on pseudo-bulked transcriptomes of each layer (pericardial, myocardial, and endocardial) across all samples, compared to the remaining layers. Pathways are grouped into (a) immune and inflammatory, (b) signaling and growth, (c) metabolic, and (d) stress- and damage-associated categories. Values represent normalized enrichment scores (NES), with positive values indicating pathway enrichment and negative values indicating depletion. Significant regulation (FDR-adjusted q-value < 0.05) is denoted by an asterisk (*).

Together, these findings outline a molecularly, potentially layered architecture of the GCM heart, in which epicardial inflammation, myocardial metabolic remodeling, and endocardial stress signatures may coexist along a continuous spatial axis.

### High-resolution spatial gene clustering across cardiac tissue layers

To achieve cell-type–resolved spatial mapping of the cardiac wall, we performed deconvolution of Stereo-seq data using the robust cell type decomposition (RCTD) algorithm^27^ with a single-cell RNA-seq reference atlas^28^. This approach enabled precise assignment of cell identities to spatial coordinates and revealed characteristic layer-specific distributions of major cardiac and immune cell types (Fig. 4a–c). Cardiomyocytes predominated across the myocardial wall, whereas fibroblasts, endothelial, and immune cells localized preferentially toward the pericardial and endocardial regions. Spatial gradients were evident for myeloid and lymphoid populations, aligning with regions of inflammatory infiltration.

**Figure 4:**
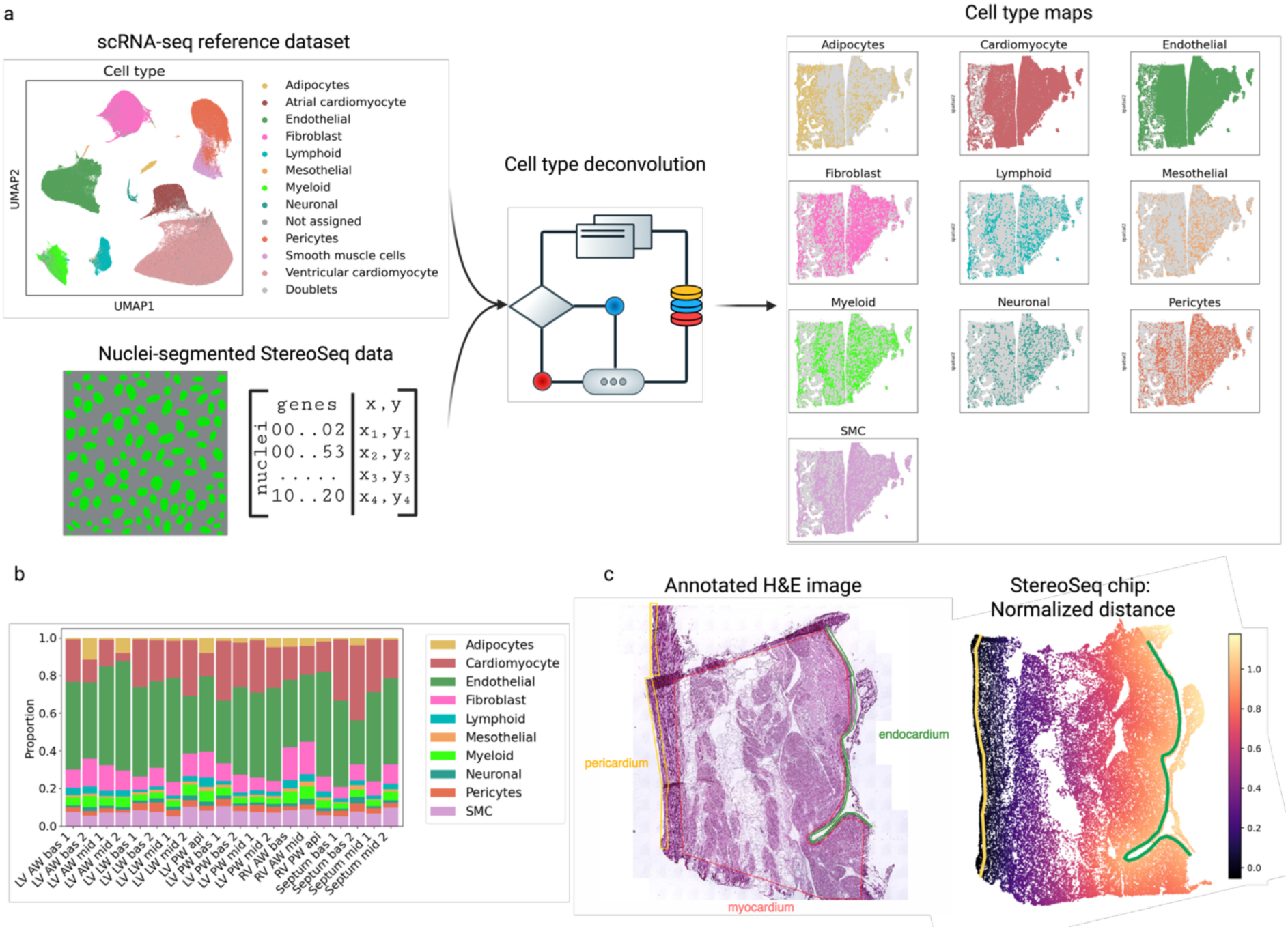
Cell-type-resolved spatial transcriptomics by robust cell type decomposition (RCTD)-based deconvolution using a single-cell RNA-seq reference atlas. (a) Uniform Manifold Approximation and Projection (UMAP) representation of the scRNA-seq reference dataset (regenerated from Litviňuková et al., Nature 2020^28^) and example of cell-type maps obtained after deconvolution with RCTD. (b) Cell-type abundances across all samples. (c) Annotation of cardiac wall layers on H&E-stained sections and corresponding annotation transfer onto the Stereo-seq chips.

To further resolve functional specialization across cardiac wall layers, we analyzed spatial distributions of cell types, gene expression, and pathway activity along the epicardial–endocardial axis. This revealed coordinated gradients of inflammatory, metabolic, and stress-response programs (Fig. 5a–g). Inflammatory signaling, including *IL-6/JAK/STAT3*, *TNF-α/NF-κB*, and interferon-response pathways, was most pronounced in the pericardial compartment, consistent with active immune infiltration and cytokine signaling near the epicardial surface. In contrast, myocardial regions exhibited relative enrichment of oxidative phosphorylation and myogenesis gene sets, compatible with preserved metabolic and contractile programs in less inflamed tissue layers. Stress- and damage-associated pathways such as apoptosis, hypoxia, and p53 pathway showed a graded increase toward the endocardium, reflecting higher levels of cellular stress in the myocardial layer.

**Figure 5:**
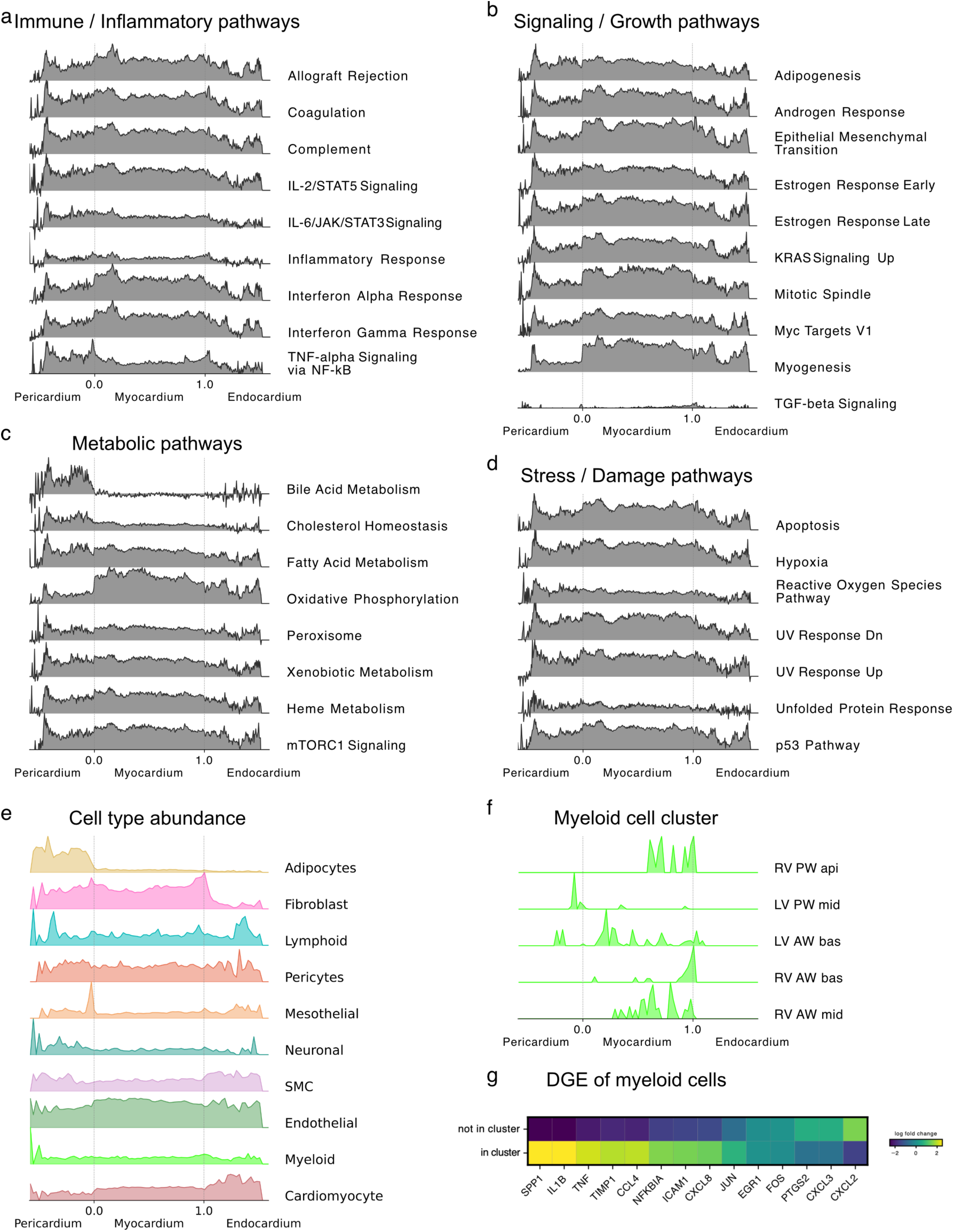
Spatial distributions of cell types, genes, and pathways along the epicardial–endocardial axis. Pathways are grouped into (a) immune and inflammatory, (b) signaling and growth, (c) metabolic, and (d) stress- and damage-associated categories. (e) Spatial distribution of cell-type abundances across the cardiac wall. (f) Spatial organization of myeloid cell clusters. (g) Differential gene expression of myeloid cells occurring within spatially defined clusters.

Differential gene expression analysis suggests that spatially clustered myeloid cells displayed a distinct transcriptional signature compared with non-clustered myeloid cells (Fig. 5g). Clustered myeloid populations were enriched for pro-inflammatory mediators, including *SPP1*, *IL1B*, *TNF*, *CXCL9*, *CXCL10*, and *CXCL12*, consistent with an activated, cytokine-secreting phenotype^29,30^. Spatial mapping of these clusters (Fig. 5f) suggests their prevalent distribution toward the pericardial and myocardial compartment, partially aligning with regional enrichment of inflammatory pathway activity. Together, these data may reflect localized gradients of myeloid activation within a compartmentalized inflammatory architecture of GCM, with focal epicardial and myocardial hotspots of immune activity embedded within a gradient of myocardial stress and remodeling.

## Discussion

IMPS represent a heterogeneous group of inflammatory cardiac diseases with diverse etiologies and clinical trajectories, ranging from self-limiting viral inflammation to rapidly progressive, immune-mediated destruction of the myocardium^3,31^. GCM constitutes the most fulminant form of this spectrum, characterized by aggressive immune infiltration, widespread myocyte necrosis, and rapid progression to end-stage heart failure^1–3^. Despite its clinical severity, the spatial organization of inflammation and tissue remodeling in IMPS and GCM remains incompletely understood. In particular, how immune activation and cardiomyocyte injury distribute across the cardiac wall has remained speculative.

Here, we applied full-transcriptome spatially resolved RNA sequencing to a human GCM explant, providing a high-resolution molecular atlas of inflammatory cardiomyopathy at near-cellular spatial resolution. Through integration of 20 samples covering 12 cardiac localizations, we characterized the cellular and transcriptional architecture of the inflamed heart wall. The spatial profiles suggested a layered organization of gene expression and immune activity, with inflammatory signaling more pronounced near the epicardium and metabolic remodeling and stress-associated programs appearing toward the myocardium and endocardium.

Across all regions, spatial clustering identified the major cardiac cell populations-cardiomyocytes, fibroblasts, endothelial, and immune cells-distributed in distinct anatomical patterns. Cardiomyocytes exhibited transcriptional diversity consistent with structural and metabolic adaptation, including upregulation of sarcomeric and mitochondrial gene programs in subsets compatible with compensatory hypertrophy, alongside a population with elevated *NPPA* and *NPPB* expression suggestive of stress activation^16,17^.

Layer-resolved pathway analysis may further imply a coordinated molecular specialization along the epicardial–endocardial axis. The pericardium displayed strong enrichment of IL-6/JAK/STAT3, TNF-α/NF-κB, and interferon-response signaling, suggesting increased inflammatory pathway engagement at the serosal interface. This inflammatory signature appeared progressively attenuated within the myocardium, where oxidative phosphorylation and myogenesis pathways dominated, consistent with preserved metabolic function and structural remodeling in surviving cardiomyocytes. In the endocardial layer, activation of apoptosis, p53, hypoxia, and unfolded-protein-response pathways could have reflected heightened cellular stress in deeper myocardial regions. Together, these data could be indicative of a layered immune–metabolic gradient, which is not directly mirrored in histopathologic or radiographic observations of GCM, where immune cell aggregates and necrosis typically distribute multifocally across all three cardiac layers^1,32–34^.

Within the immune compartment, spatial mapping and deconvolution may point to a pronounced heterogeneity among myeloid and lymphoid cell populations. Myeloid cells formed discrete spatial clusters enriched for pro-inflammatory mediators, including *SPP1*, *IL1B*, *TNF*, *CXCL9*, *CXCL10*, and *CXCL12*, consistent with highly activated, cytokine-secreting macrophage subsets^29,30^. These clusters distributed preferentially toward the pericardial compartment, in line with the enrichment of inflammatory pathways in that region. In contrast, more diffuse myeloid populations in the myocardium exhibited lower expression of inflammatory genes, potentially reflecting partial transition toward tissue-remodeling phenotypes. B cells organized into two transcriptionally distinct lymphoid clusters: an *IGHM*^high^, *JCHAIN*^+^ subset corresponding to early or naïve B cells, and an *IGHG4*^high^ population representing class-switched, antibody-secreting plasma cells^20,21,23–25^. The coexistence of IgM- and IgG4-producing populations within the inflamed cardiac wall may indicate ongoing B cell maturation and sustained humoral activity, paralleling features of chronic autoimmune inflammation^35–37^.

The predominance of IL-6–related signaling and cytokine-producing myeloid cells in the pericardial region raises the possibility that immune activation in GCM may originate near the epicardial surface and propagate inward-a mechanism conceptually analogous to the “outside-in” inflammatory trajectory observed in Dressler syndrome and other autoimmune serositis syndromes^38–40^. While speculative, this model provides a spatial framework compatible with the hypothesis that pericardial immune activation could precede and drive myocardial injury in IMPS. The observed enrichment of IL-6/JAK/STAT3 signaling is of particular relevance given the emerging interest in IL-6 receptor blockade in inflammatory cardiomyopathies and myocarditis-associated heart failure^41^.

Within the myocardium, metabolic remodeling and sarcomeric reprogramming may reflect adaptive responses of surviving cardiomyocytes under sustained cytokine stress. The coordinated upregulation of oxidative phosphorylation and *MYC* target pathways could represent compensatory increases in mitochondrial activity to meet increased energetic demands, consistent with adaptive hypertrophy. The endocardial enrichment of stress and damage-associated pathways, including apoptosis and hypoxia signaling, may reflect downstream consequences of impaired perfusion and sustained inflammation, akin to the transition from active myocarditis to fibrotic end-stage remodeling observed in chronic inflammatory cardiomyopathies.

These findings align with and extend recent spatial transcriptomic studies of the healthy and failing human heart, which have demonstrated distinct immune niches and stromal–immune cell interactions^42,43^. In particular, the localization of *SPP1*^high^ myeloid cells adjacent to fibroblast-rich regions mirrors macrophage–fibroblast communication axes implicated in fibrotic remodeling^44,45^. The identification of *IGHG4*^+^ plasma cells and myeloid clusters co-localized within inflamed tissue potentially reflects the presence of tertiary lymphoid–like structures, echoing findings from chronic autoimmune myocarditis and systemic inflammatory disorders^46,47^.

A key strength of this study is the integration of high-density spatial transcriptomic data with anatomical annotation, capturing the full transmural architecture of the human cardiac wall across multiple ventricular levels. This design allowed systematic comparison of transcriptional programs within and across layers, revealing recurrent spatial gradients despite regional heterogeneity. Although the analyses are derived from a single explanted heart, the consistency of layer-associated signatures in 20 samples across 12 localizations supports their biological plausibility. One important consideration when interpreting these findings is that the transcriptional profiles reflect disease under intensified immunosuppressive therapy (see Supplementary Note: *Clinical course of the patient with GCM*). In particular, the patient received escalated immunosuppression immediately prior to transplantation, which may have influenced immune cell activation states and cellular phenotypes in the explanted tissue. Moreover, the transcriptional changes observed in cardiomyocyte subpopulations, especially signatures of remodeling and adaptive hypertrophy, may not solely reflect inflammation-driven responses but also profound adaptations to end-stage heart failure and the influence of inotropic therapy.

Together, these observations refine current perspectives on the spatial organization of inflammation and remodeling in GCM. They outline a spatial framework in which epicardial-skewed immune activation, myocardial metabolic adaptation and injury, and endocardial stress may appear sequentially arranged along the cardiac wall. While mechanistic inferences remain limited, this layered architecture could be compatible with a model of myocarditis in which early cytokine signaling and localized immune activation at the pericardial interface contribute to broader myocardial involvement. From a translational perspective, these findings highlight the potential relevance of targeting early cytokine and myeloid activation-particularly the IL-6 axis-in inflammatory cardiomyopathies.

In summary, spatially resolved transcriptomics of a human GCM explant suggest a layered immune–metabolic architecture across the cardiac wall, with inflammatory signaling more prominent near the epicardium and metabolic as well as stress-associated programs observed toward the myocardium and endocardium. These data offer a molecular reference for the spatial organization of inflammation in fulminant autoimmune myocarditis and may provide a reference for future spatial analyses of inflammatory cardiomyopathies and therapeutic interventions.

## Online methods

### Heart explantation, dissection & sample pre-processing

The explanted heart was obtained from a patient undergoing orthotopic heart transplantation for end-stage GCM. Clinical details and the perioperative course of this patient are described in the supplementary note. Immediately after cardioplegic arrest, the organ was excised following standard surgical procedures. On a sterile back table in the operating room, the explanted heart was immediately flushed with cold isotonic sodium chloride solution to remove residual blood and was then dissected into several predefined blocks corresponding to distinct regions of interest.

In total, 20 tissue blocks from defined cardiac regions-including septal, right and left ventricular anterior, lateral, and posterior walls-were sectioned intraoperatively. The samples were transferred into cryotubes and immediately snap-frozen in liquid nitrogen. All specimens were subsequently transported to the research laboratory and stored at -80 °C until further processing. For further subdivision, the frozen tissue blocks were trimmed using a high-frequency oscillating saw under continuous cooling to prevent thawing and preserve RNA integrity. Tissue pieces measuring approximately 1-2 cm x 1-2 cm were embedded in cryomolds using optimal cutting temperature (OCT) compound.

Prior to Stereo-seq experiments, total RNA integrity (RIN) was assessed using a homogenized small heart tissue sample of the donated GCM heart. RNA was isolated and purified (Qiagen, RNeasy Mini Kit) and RNA integrity was measured utilizing a tape station, yielding a RIN of 7.8.

### Stereo-seq wet lab

Fresh-frozen tissue blocks were processed following the manufacturer’s protocol (S*tereo-seq Transcriptomics Set for Chip-on-a-Slide User Manual Ver. B*). Tissue sections of 10 µm thickness were prepared using a cryostat and mounted onto Stereo-seq chips (Kit Ver. 1.2, STOmics, China). To accommodate the 1 x 1 cm chip format, tissue blocks exceeding 1 cm in any dimension were divided and mounted onto separate chips. Mounted sections were fixed in methanol at –20 °C and subsequently stained with FITC-labelled ssDNA. Tissue sections and fiducial grid lines were imaged with a widefield fluorescent microscope (Metasystems, Germany) equipped with a 10x objective. Image quality was assessed using imageQC software (STOmics, China). After imaging, tissue sections were permeabilized for 12 minutes to enable RNA release, as determined in a prior permeabilization optimization experiment (*Stereo-seq Permeabilization Set for Chip-on-a-Slide User Manual Ver. B*). RNA was captured by spatially barcoded DNA nanoballs and reverse transcribed into cDNA. The spatially barcoded cDNA was released, purified, and amplified by PCR, followed by fragmentation. Sequencing libraries were constructed according to the manufacturer’s protocol and sequenced on the DNBSEQ platform (MGI, China) at STOmics Riga.

For each sample, an adjacent tissue section was mounted on a SuperFrost Plus microscope slide and stained with hematoxylin and eosin (H&E). Images of H&E-stained sections were acquired using a widefield bright-field microscope (Metasystems, Germany) equipped with an RGB light source.

### Stereo-seq data management & analysis

#### Preprocessing

Genome alignment of raw sequencing data and image-to-chip registration were performed by STOmics using SAW software V8^48^ (STOmics, China), yielding gene expression files (.gef) for each tissue section. For downstream analysis, cell-binned and bin100 data were imported into anndata objects using the Stereopy (v.1.6.0) toolkit^49^.

### Bin100

Anndata objects from all samples were loaded, concatenated, and filtered using the following quality thresholds: minimum counts per bin, 500; maximum counts per bin, 20,000; minimum genes per bin, 250; minimum bins per gene, 100. Raw count data were normalized using Pearson residuals (*sc.experimental.pp.normalize_pearson_residuals,* default parameters) as implemented in Scanpy (v.1.9.1)^50^, which stabilizes variance by modeling counts under a negative binomial distribution. All remaining genes were retained for principal component analysis (PCA), and the first 30 principal components were used for downstream analysis.

To correct for sample-specific batch effects, principal components were harmonized using the rapids-singlecell (v.0.3.1) implementation^51^ of Harmony^52^ (θ = 2, default parameters), which converged after 10 iterations. The corrected principal components were used to construct a latent-space neighborhood graph. Leveraging spatial coordinates, a spatial graph for each sample was generated using Squidpy (v. 1.2.4)^53^ with 10 nearest neighbours. Both graphs were combined to perform spatially aware clustering with SpatialLeiden (v.0.2.0)^54^ (latent-space resolution = 0.7; spatial resolution = 1.0; layer ratio = 1.0).

Differential gene expression analysis among the resulting 15 clusters was performed using the Wilcoxon rank-sum test as implemented in Scanpy, with p-values adjusted for multiple testing using the Benjamini–Hochberg procedure. Clusters were annotated based on the top 50 differentially expressed genes indicating the dominant cell type per bin (see Supplementary Table). The distribution of annotated cell types was visualized using UMAP embeddings of the latent-space graph and corresponding spatial scatter plots.

The myeloid subcluster was binarily classified into active bins with high or low inflammatory activation. We calculated a gene module-score for a gene set indicative of inflammatory activation (*IL1B, TNF, NFKBIA, CXCL2, CXCL3, CXCL8, CCL2, CCL3, CCL4, PTGS2, ICAM1, EGR1, FOS, JUN, SPP1, TIMP1*) using *Scanpy*’s score_genes function with log-normalized data. We classified cells into two categories: high inflammatory activation and low inflammatory activation using the median score (50^th^ percentile) within the subset as a threshold. This binary label was used for downstream analyses and visualization.

Anatomical regions corresponding to cardiac wall layers (pericardium, myocardium, and endocardium) were manually annotated in adjacent H&E-stained images. The H&E imagesand their annotation masks were registered to the DAPI-stained Stereo-seq images using a Python implementation of the thin-plate spline transformation^55^ applied on manually defined paired landmarks across image modalities, followed by manual adjustment if required.

Gene set enrichment analysis (GSEA) was performed on pseudo-bulk expression profiles aggregated by anatomical region (pericardium, myocardium, and endocardium). For each layer, genes were ranked according to the difference in mean expression between that region and the remaining layers. Pre-ranked GSEA was conducted using GSEApy (v.1.1.10)^56^ with the MSigDB Hallmark 2020 gene set collection^26^, a minimum gene set size of 15, a maximum of 500, and 1,000 permutations. Normalized enrichment scores (NES) and FDR-adjusted q-values were reported for each pathway. Pathways with an FDR-adjusted q-value < 0.05 were grouped into functional categories and visualized using heatmaps.

### Cell bin

Cell-type annotation of cell-binned Stereo-seq data was performed using robust cell type decomposition (RCTD, spacxr v.2.2.1)^27^, which infers the most probable cell type for each spatial bin by modeling observed transcript counts as mixtures of single-cell reference profiles. We used a publicly available single-cell RNA-sequencing atlas of the human heart^28^ as the reference dataset. The reference dataset was loaded via SeuratDisk^57^, and raw gene expression counts together with cell-type labels were extracted to create the RCTD reference object. For each sample, the Stereo-seq data were converted into SingleCellExperiment^58^ objects using zellkonverter^59^, from which the gene expression matrices and spatial coordinates were extracted to generate spatial-RNA objects for RCTD. The reference and spatial-RNA objects were combined to run RCTD in doublet mode (min_UMI = 10) to account for potential mixed or overlapping bins. RCTD internally normalizes sequencing depth and platform-specific differences using a Poisson-based model. The resulting singlet and doublet predictions provided spatially resolved cell-type assignments across each section.

To validate the RCTD-derived annotations, we performed differential gene expression (DGE) analysis on the predicted cell types using GPU-accelerated logistic regression as implemented in rapids-singlecell (v.0.3.1), applied to normalized-to-median and log1p-transformed counts. For anatomical regions in which the peri-, myo-, and endocardium were captured on one or multiple Stereo-seq chips, spatial gradient analyses were performed. Locations covered by multiple chips were merged into a single object. Based on heart layer annotations (see “Methods: bin100”), baseline contours were defined for the pericardial and endocardial layers. To quantify spatial positioning relative to these anatomical boundaries, signed Euclidian distances were computed from each cell to the nearest point on the pericardial and endocardial baselines. The baseline contours were sampled every 10 μm, and the nearest segment for each cell was identified using a *k*-d tree nearest-neighbor search. Signed distances were calculated using the 2D cross product to encode directionality (negative values inside, positive values outside). For closed polygons, distances were assigned negative values for cells located within the polygon and positive values otherwise.

Distances into pericardial (d_peri_) and endocardial (d_endo_) boundaries were combined into a normalized gradient metric:

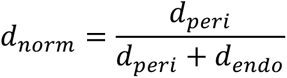

This approach captured the continuous spatial axis between the pericardium and endocardium. Along these axes, ridge plots were generated to visualize the distributions of cell- or gene-level-features. Cells were binned along the normalized distance axis (default: n = 500 bins), and for each group, either the fraction of cells or the mean feature value per bin was computed. The resulting bin-wise distributions were scaled within groups and plotted, optionally separated by sample or feature.

## Ethics

This study was approved by the ethics committee of Charité – Universitätsmedizin Berlin (EA4/163/21), and the patient provided informed written consent on participating in this study.

## Supporting information

Supplementary note: Clinical course of the patient with GCM

## Data Availability

All data produced in the present study are available upon reasonable request to the authors.

## Acknowledgements

We thank the patient for her generous contribution of cardiac tissue, which made this study possible.

Nicolas Musigk was supported by a stipend from the Deutsche Herzstiftung e.V.. Bettina Heidecker is funded by the Deutsches Herzzentrum Berlin (DHZB) Foundation and was participant in the BIH-Charité Advanced Clinician Scientist Pilotprogram funded by the Charité –Universitätsmedizin Berlin and the Berlin Institute of Health. We acknowledge the Scientific Computing of the IT Division at Charité - Universitätsmedizin Berlin for providing computational resources that supported the analyses reported in this work. We further thank Charité’s Research Data Management service for providing and maintaining the OMERO platform, and the iPATH.Berlin - Immunopathology for Experimental Models core unit at Charité –Universitätsmedizin Berlin for performing the H&E staining. We also thank Ines Koch from the Institute of Pathology at Charité – Universitätsmedizin Berlin for her assistance with the sectioning of frozen OCT-embedded samples.

## References

1. Kociol, R.D., et al. Recognition and Initial Management of Fulminant Myocarditis: A Scientific Statement From the American Heart Association. Circulation 141, e69–e92 (2020).

2. Drazner, M.H., et al. 2024 ACC Expert Consensus Decision Pathway on Strategies and Criteria for the Diagnosis and Management of Myocarditis. JACC 85, 391–431 (2025).

3. Schulz-Menger, J., et al. 2025 ESC Guidelines for the management of myocarditis and pericarditis: Developed by the task force for the management of myocarditis and pericarditis of the European Society of Cardiology (ESC)Endorsed by the Association for European Paediatric and Congenital Cardiology (AEPC) and the European Association for Cardio-Thoracic Surgery (EACTS). European Heart Journal (2025).

4. Antony, V.B. Immunological mechanisms in pleural disease. Eur Respir J 21, 539–544 (2003).

5. Møllgård, K., et al. A mesothelium divides the subarachnoid space into functional compartments. Science 379, 84–88 (2023).

6. Theis, J.L., et al. Expression patterns of cardiac myofilament proteins: genomic and protein analysis of surgical myectomy tissue from patients with obstructive hypertrophic cardiomyopathy. Circ Heart Fail 2, 325–333 (2009).

7. Sheikh, F., Lyon, R.C. & Chen, J. Functions of myosin light chain-2 (MYL2) in cardiac muscle and disease. Gene 569, 14–20 (2015).

8. Branco, A.F., et al. Gene Expression Profiling of H9c2 Myoblast Diaerentiation towards a Cardiac-Like Phenotype. PLoS One 10, e0129303 (2015).

9. Vega, R.B., Horton, J.L. & Kelly, D.P. Maintaining ancient organelles: mitochondrial biogenesis and maturation. Circ Res 116, 1820–1834 (2015).

10. Moore, J., et al. Multi-Omics Profiling of Hypertrophic Cardiomyopathy Reveals Altered Mechanisms in Mitochondrial Dynamics and Excitation-Contraction Coupling. Int J Mol Sci 24(2023).

11. Witjas-Paalberends, E.R., et al. Mutations in MYH7 reduce the force generating capacity of sarcomeres in human familial hypertrophic cardiomyopathy. Cardiovasc Res 99, 432–441 (2013).

12. Tan, Z., et al. Systemic Bioinformatic Analyses of Nuclear-Encoded Mitochondrial Genes in Hypertrophic Cardiomyopathy. Front Genet 12, 670787 (2021).

13. Hu, H., et al. Long noncoding RNA MALAT1 enhances the apoptosis of cardiomyocytes through autophagy modulation. Biochem Cell Biol 98, 130–136 (2020).

14. Nomura, S., et al. Cardiomyocyte gene programs encoding morphological and functional signatures in cardiac hypertrophy and failure. Nat Commun 9, 4435 (2018).

15. Huang, S., et al. Long noncoding RNA MALAT1 mediates cardiac fibrosis in experimental postinfarct myocardium mice model. J Cell Physiol 234, 2997–3006 (2019).

16. Sergeeva, I.A., et al. Identification of a regulatory domain controlling the Nppa-Nppb gene cluster during heart development and stress. Development 143, 2135–2146 (2016).

17. Goetze, J.P., et al. Cardiac natriuretic peptides. Nat Rev Cardiol 17, 698–717 (2020).

18. Zarate, Y.A., et al. ACTC1 Variants Result in Isolated and Syndromic Cardiac Phenotypes. Clin Genet (2025).

19. Vigil-Garcia, M., et al. Gene expression profiling of hypertrophic cardiomyocytes identifies new players in pathological remodelling. Cardiovasc Res 117, 1532–1545 (2021).

20. Mattoo, H., et al. De novo oligoclonal expansions of circulating plasmablasts in active and relapsing IgG4-related disease. J Allergy Clin Immunol 134, 679–687 (2014).

21. Kamisawa, T., Zen, Y., Pillai, S. & Stone, J.H. IgG4-related disease. Lancet 385, 1460–1471 (2015).

22. Hsieh, S.C., et al. The Cellular and Molecular Bases of Allergy, Inflammation and Tissue Fibrosis in Patients with IgG4-related Disease. Int J Mol Sci 21(2020).

23. Delves, P.J. & Roitt, I.M. The Immune System. New England Journal of Medicine 343, 37–49 (2000).

24. Shlomchik, M.J. & Weisel, F. Germinal center selection and the development of memory B and plasma cells. Immunol Rev 247, 52–63 (2012).

25. Randall, T.D., Brewer, J.W. & Corley, R.B. Direct evidence that J chain regulates the polymeric structure of IgM in antibody-secreting B cells. J Biol Chem 267, 18002–18007 (1992).

26. Liberzon, A., et al. The molecular signatures database hallmark gene set collection. Cell systems 1, 417–425 (2015).

27. Cable, D.M., et al. Robust decomposition of cell type mixtures in spatial transcriptomics. Nature biotechnology 40, 517–526 (2022).

28. Litviňuková, M., et al. Cells of the adult human heart. Nature 588, 466–472 (2020).

29. Fajgenbaum, D.C. & June, C.H. Cytokine Storm. New England Journal of Medicine 383, 2255–2273 (2020).

30. Shirakawa, K., et al. Interleukin-7 Receptor Activation in Interstitial Macrophages Promotes Lung Fibrosis through Spp1. Am J Respir Cell Mol Biol (2025).

31. Musigk, N., et al. The inflammatory spectrum of cardiomyopathies. Frontiers in Cardiovascular Medicine 11(2024).

32. Litovsky, S.H., Burke, A.P. & Virmani, R. Giant cell myocarditis: an entity distinct from sarcoidosis characterized by multiphasic myocyte destruction by cytotoxic T cells and histiocytic giant cells. Mod Pathol 9, 1126–1134 (1996).

33. Basso, C. Myocarditis. New England Journal of Medicine 387, 1488–1500 (2022).

34. Li, J.H., et al. Subendocardial Involvement as an Underrecognized Cardiac MRI Phenotype in Myocarditis. Radiology 302, 61–69 (2022).

35. Lighaam, L.C. & Rispens, T. The Immunobiology of Immunoglobulin G4. Semin Liver Dis 36, 200–215 (2016).

36. Motta, R.V. & Culver, E.L. IgG4 autoantibodies and autoantigens in the context of IgG4-autoimmune disease and IgG4-related disease. Front Immunol 15, 1272084 (2024).

37. Zografou, C., Vakrakou, A.G. & Stathopoulos, P. Short- and Long-Lived Autoantibody-Secreting Cells in Autoimmune Neurological Disorders. Front Immunol 12, 686466 (2021).

38. Cremer, P.C., Klein, A.L. & Imazio, M. Diagnosis, Risk Stratification, and Treatment of Pericarditis: A Review. JAMA 332, 1090–1100 (2024).

39. Kahlenberg, J.M. & Kang, I. Advances in Disease Mechanisms and Translational Technologies: Clinicopathologic Significance of Inflammasome Activation in Autoimmune Diseases. Arthritis Rheumatol 72, 386–395 (2020).

40. Wahren-Herlenius, M. & Dörner, T. Immunopathogenic mechanisms of systemic autoimmune disease. Lancet 382, 819–831 (2013).

41. Savvatis, K., et al. Interleukin-6 receptor inhibition modulates the immune reaction and restores titin phosphorylation in experimental myocarditis. Basic Res Cardiol 109, 449 (2014).

42. Kanemaru, K., et al. Spatially resolved multiomics of human cardiac niches. Nature 619, 801–810 (2023).

43. Amrute, J.M., et al. Targeting immune-fibroblast cell communication in heart failure. Nature 635, 423–433 (2024).

44. Hoeft, K., et al. Platelet-instructed SPP1(+) macrophages drive myofibroblast activation in fibrosis in a CXCL4-dependent manner. Cell Rep 42, 112131 (2023).

45. Uhlig, M., Billig, S., Wienhold, J. & Schumacher, D. Pro-Fibrotic Macrophage Subtypes: SPP1+ Macrophages as a Key Player and Therapeutic Target in Cardiac Fibrosis? Cells 14(2025).

46. Dong, Y., Wang, T. & Wu, H. Tertiary lymphoid structures in autoimmune diseases. Front Immunol 14, 1322035 (2023).

47. Zhu, M., et al. Cardiac ectopic lymphoid follicle formation in viral myocarditis involving the regulation of podoplanin in Th17 cell diaerentiation. Faseb j 35, e21975 (2021).

48. Gong, C., et al. SAW: an eaicient and accurate data analysis workflow for Stereo-seq spatial transcriptomics. GigaByte 2024, gigabyte111 (2024).

49. Fang, S., et al. Stereopy: modeling comparative and spatiotemporal cellular heterogeneity via multi-sample spatial transcriptomics. Nat Commun 16, 3741 (2025).

50. Wolf, F.A., Angerer, P. & Theis, F.J. SCANPY: large-scale single-cell gene expression data analysis. Genome Biol 19, 15 (2018).

51. Avantika, L.C., Ilango, R. & Dicks, S. NVIDIA-Genomics-Research/rapids-single-cell-examples: v2022. 12.0 (v2022. 12.0). Zenodo, European Organization for Nuclear Research (CERN) (2023).

52. Korsunsky, I., et al. Fast, sensitive and accurate integration of single-cell data with Harmony. Nat Methods 16, 1289–1296 (2019).

53. Palla, G., et al. Squidpy: a scalable framework for spatial omics analysis. Nature methods 19, 171–178 (2022).

54. Müller-Bötticher, N., Sahay, S., Eils, R. & Ishaque, N. SpatialLeiden: spatially aware Leiden clustering. Genome Biology 26, 24 (2025).

55. Bookstein, F.L. Principal warps: Thin-plate splines and the decomposition of deformations. IEEE Transactions on pattern analysis and machine intelligence 11, 567–585 (2002).

56. Fang, Z., Liu, X. & Peltz, G. GSEApy: a comprehensive package for performing gene set enrichment analysis in Python. Bioinformatics 39, btac757 (2023).

57. Hoaman, P. SeuratDisk: Interfaces for HDF5-Based Single Cell File Formats. (2023).

58. Amezquita, R.A., et al. Orchestrating single-cell analysis with Bioconductor. Nature Methods 17, 137–145 (2020).

59. Lun, L.Z.a.A. zellkonverter: Conversion Between scRNA-seq Objects. (2025).

