## Supplementary note: Clinical course of the patient with GCM for "Spatial transcriptomics reveals layered immune-metabolic architecture of giant cell myocarditis"

A woman in her fifties presented with progressive dyspnea and reduced exercise tolerance corresponding to New York Heart Association class III heart failure.

Echocardiographic parameters demonstrated marked left ventricular dilation (LVIDd = 5.98 cm), wall thinning (IVSd = 1.07 cm, LVPWd = 0.88 cm), and globally reduced longitudinal strain (GLS = –9.8%), consistent with severe systolic dysfunction and advanced myocardial remodeling (Supplementary Fig. c–e). Cardiac magnetic resonance imaging revealed late gadolinium enhancement predominantly involving the lateral and apical segments of the left ventricle and T2-weighted myocardial edema, consistent with active myocarditis (Supplementary Fig. a,b). Endomyocardial biopsy confirmed extensive myocyte necrosis with multinucleated giant cells, establishing the diagnosis of GCM.

Despite immunosuppressive therapy with prednisolone and azathioprine, later intensified mycophenolate mofetil and tacrolimus, the patient developed progressive biventricular failure, pulmonary hypertension, and acute kidney injury. Repeated biopsies demonstrated ongoing inflammation with persistent giant cells and increasing fibrosis, indicating refractory disease.

Two years after diagnosis, the patient underwent orthotopic heart transplantation for end-stage heart failure. Immediately before transplantation, the patient required milrinone for inotropic support, and an intensified immunosuppressive regimen with prednisolone and tacrolimus was initiated as part of the pre-transplant protocol, which may have influenced the immunologic and transcriptional profile of the explanted heart. The explanted organ was used for the present spatial transcriptomic and histologic analyses.

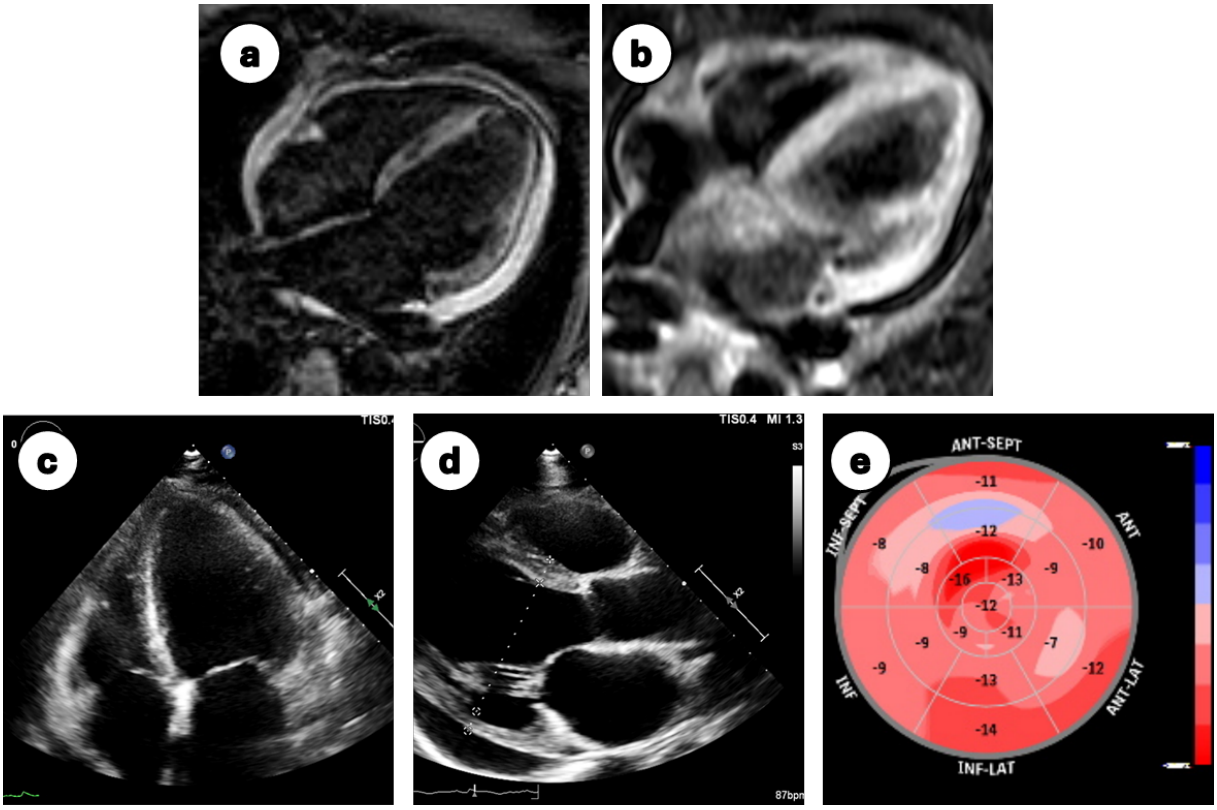


***Supplementary figure: Cardiac imaging at the time of diagnosis.*** *(a,b) Cardiac magnetic resonance imaging showing typical features of active myocarditis. (a) Late gadolinium enhancement (LGE) predominantly affecting the lateral and apical segments of the left ventricle. (b) Corresponding myocardial edema on T2-weighted imaging. (c,d) Transthoracic echocardiography revealing a markedly dilated left ventricle in the apical four-chamber (c) and parasternal long-axis (d) views (interventricular septal diameter [IVSd], 1.07 cm; left ventricular internal diameter in diastole [LVIDd], 5.98 cm; left ventricular posterior wall diameter [LVPWd], 0.88 cm). (e) Global longitudinal strain map demonstrating globally reduced strain values (average −9.8%) with an estimated left ventricular ejection fraction of approximately 35%.*
